# Mechanical thrombectomy with the Vecta 46 catheter: a safety and outcome analysis

**DOI:** 10.1101/2025.10.17.25338262

**Authors:** Hunter Hutchinson, Chloe DeYoung, Danyas Sarathy, Grace Hey, Wiley Gillam, Shawna Amini, Muhammad Abdul Baker Chowdhury, Brandon Lucke-Wold, Zachary Sorrentino, Matthew Koch

**Affiliations:** College of Medicine, University of Florida, Gainesville, FL 32610, USA; Lillian S. Wells Department of Neurosurgery, University of Florida, Gainesville, FL 32610, USA

## Abstract

2.

**Background:** The AXS Vecta 46 intermediate catheter (Stryker) provides a large enough inner diameter to achieve effective aspiration with a small enough outer diameter and soft distal-tip flexibility to track safely through more tortuous and smaller arterial segments to non-traumatically target medium vessel occlusions. The efficacy of the Vecta 46 in the spectrum of large and medium vessel occlusions has not been well elucidated in the literature.

**Methods:** This retrospective cohort study included patients who underwent MT for acute ischemic stroke at the University of Florida between July 2022 and June 2024. The outcomes of patients treated with the Vecta 46 was compared to that of all other catheters used at the institution.

**Results:** The distribution of aspiration and stent retriever attempts in Vecta 46 procedures versus non-Vecta 46 procedures was significantly different (p = 0.00325). Aspiration was attempted 1.66 ± 0.936 times in the Vecta 46 group and 1.12 ± 0.650 times in the non-Vecta 46 group (p = 0.00135). More mechanical thrombectomies with the Vecta 46 included aspiration of a secondary thrombus (p = 0.0314), despite no difference in distribution of primary or secondary occlusion location. There were no statistically significant differences in recanalization success (p = 0.800), recanalization time (p = 0.245), procedure duration (p = 0.580), discharge modified Rankin Score (p = 0.875), or intracranial hemorrhage rate (p = 0.720) between non-Vecta 46 and Vecta 46 procedures.

**Conclusions:** Vecta 46 has similar safety and functional outcomes compared to other endovascular treatment options despite procedural differences.

## 3. Introduction

Acute ischemic stroke caused by proximal large vessel occlusions (LVO) have conclusively been shown to benefit from mechanical thrombectomy (MT) over the past decade, as the result of multiple large randomized controlled trials, cementing this procedure as the standard of care for eligible LVO patients. MT for acute ischemic stroke has led to significantly improved recanalization rates and functional outcomes compared to medical therapy alone across numerous landmark clinical trials.^1–3^ These trials, however, predominately enrolled patients with proximal occlusions, such as in the internal carotid or proximal middle cerebral arteries and only had small fractions of patients with more distal occlusions. They also focused on stent-retriever devices (SR), which have since been augmented with other viable reperfusion techniques, such as direct aspiration.^4–9^ Thus, the efficacy of MT for medium vessel occlusions (MeVOs), such as the M2/3 and analogous segments in the anterior and posterior cerebral arteries, remain less certain in early trial data. In pooled subgroup analyses across multiple trials, MT for M2 occlusions showed a non-significant trend toward benefit, and thus current guidelines endorse MT in only select distal vessels with a ‘weak’ recommendation level.^10,11^

Nonetheless, MeVO strokes are increasingly recognized as clinically impactful, and growing experience in high-volume stroke centers is driving broadening of MT indications to include distal occlusions in select cases. There is emerging evidence from several retrospective studies and registries that show that MeVO MT is both technically feasible, and associated with similar recanalization rates and outcome measures as those in LVO MT.^12^ This open question is being addressed in ongoing studies such as the DISTAL and ESCAPE-MeVO trials, which reflects the growing interest of extending endovascular treatment to smaller-caliber occlusions.

The technical nuance of MeVO MT lies in the anatomy: distal intracranial vessels are narrower, more tortuous, and fragile compared to proximal vessels, increasing the difficulty of navigation and procedural complication due to vessel injury from catheter associated mechanical trauma. Despite these thrombi being smaller in size, medical therapy still fails to recanalize a substantial number of MeVO cases, which provided the impetus to create novel devices optimized for distal circulation MT to help circumvent the anatomic barrier that limits access to these MeVO. Newer generation aspiration catheters feature reduced outer diameters, and better distal flexibility, with enough lumen size to generate effective aspiration. One such catheter at use in our center is the AXS Vecta 46 Intermediate catheter (Stryker), which has an inner diameter of 0.046”, straddling the line between that of smaller bore aspiration catheters (on the order of 0.035”), and larger/super-bore catheters (frequently 0.070” – 0.088”). There are many such devices that have subsequently released on the market (such as the Penumbra 4MAX [0.041”], or Sofia 5F [0.055”]), and the general principle across all these innovations is to enable a large enough inner diameter to achieve effective aspiration, however a small enough outer diameter with soft distal-tip flexibility to track safely through more tortuous and smaller arterial segments to non-traumatically target MeVO.

Despite the availability of these newer intermediate aspiration devices, there is a notable lack of published clinical data evaluating their safety and effectiveness in routine stroke MT practice. Early experiences have been limited to small case series or conference reports focusing on distal vessel usage, and Vecta 46 catheter performance has not been well characterized in the literature.^13–15^ To address this gap, we conducted a retrospective safety and feasibility study of the Vecta 46 intermediate catheter in acute ischemic stroke MT at the University of Florida comprehensive stroke center. Our primary aim was to describe the safety profile and clinical outcomes associated with Vecta 46 use in MeVO and LVO cases, and to determine whether these outcomes are comparable to those achieved with other larger bore catheters used at our institution. Herein, we demonstrate that MT utilizing Vecta 46 is safe and efficacious for both LVO and MeVO strokes.

## 4. Methods

### 2.1 Study Design and Population

This retrospective cohort study included patients who underwent MT for acute ischemic stroke at the University of Florida between July 2022 and June 2024. A total of 174 MT procedures were reviewed. Inclusion criteria included adult patients (≥18 years old) who underwent MT. Patients with incomplete procedural or outcome data were excluded from this analysis. Patients who underwent MT with both the Vecta 46 catheter, along with other catheters were included and compared. The study was approved by the local Institutional Review Board with a waiver of informed consent.

### 2.2 Data Collection

Patient demographic information, clinical risk factors, stroke characteristics, procedural details, and outcomes were extracted from the electronic medical record. Specific demographic and clinical information collected includes age, sex, comorbidities (including atrial fibrillation, diabetes mellitus, hypertension, coronary artery disease, prior cerebrovascular events, and smoking history), baseline modified Rankin Scale (mRS) and initial stroke severity score (National Institute of Health Stroke Scale (NIHSS) score on admission. Technique used (aspiration or SR), number of aspiration and SR attempts, vessel of primary occlusion and vessel of secondary inclusion were included in procedure details. Occlusion locations included anterior cerebral artery (ACA), middle cerebral artery (MCA), internal carotid artery (ICA), common carotid artery (CCA), posterior cerebral artery (PCA), basilar artery, and vertebral artery. Segments (1, 2 or 3) of ACA and MCA occlusions were also recorded. PCA, basilar artery, and vertebral artery occlusions were grouped as “posterior circulation.”

### 2.3 Outcomes

The primary measure was the rate of successful revascularization, which we defined using the Thrombolysis in Cerebral Infarction (TICI) scale. A successful revascularization was defined as TICI 2b or greater. Procedure outcomes included time to recanalization, procedure duration, modified Rankin Scale (mRS) at discharge and intracranial hemorrhage (ICH) incidence. Follow-up was defined as any documented outpatient neurology or stroke clinic visit where mRS was reassessed, with time from discharge to follow-up measured in days.

### 2.4 Statistical Analysis

Comparisons between Vecta 46 and non-Vecta MTs were made using chi-square for categorical variables, Student’s t-tests for parametric numerical variables, and Mann-Whitney U tests for non-parametric numerical variables. Distribution of data was determined with the Shapiro-Wilk test. A p-value < 0.05 was defined as statistically significant. All clinical data were extracted electronic medical records and analyzed in R (R Core Team 2025. Vienna, Austria: R Foundation for Statistical Computing).

## 5. Results

### 3.1 Baseline Characteristics of Patients

A total of 174 patient MTs were analyzed. Vecta 46 catheters were used in 32 MTs and other catheters were used in 142 MTs. Baseline characteristics of the patients treated with the Vecta 46 or other catheters were not significantly different. The mean age of the non-Vecta 46 cohort was 69.4 ± 14.9 years old and 68.7 ± 17.8 years old (p = 0.537). There were 77 (54.2%) male and 65 (46.5%) female patients in the non-Vecta 46 group. There were 16 (50%) male and 16 (50%) female patients in the Vecta 46 group. The non-Vecta 46 group included 37 (26.1%) patients with atrial fibrillation, 21 (14.7%) patients with diabetes mellitus, 94 (66.2%) with hypertension, 39 (27.5%) patients with coronary artery disease (CAD), 25 (17.6%) patients with a history of cerebrovascular accident/transient ischemic attack (CVA/TIA), and 44 (31.0%) patients were current smokers. The Vecta 46 group included 4 (12.5%) patients with atrial fibrillation, 7 (21.9%) patients with diabetes mellitus, 22 (69.0%) patients with hypertension, 6 (18.8%) patients with CAD, 5 (15.6%) patients with a history CVA/TIA, and 6 (18.8%) patients were current smokers. The mean baseline mRS was 0.570 ± 1.01 in the non-Vecta 46 group and 0.81 ± 1.62 in the Vecta 46 group. The mean admission NIHSS was 17.2 ± 7.66 in the non-Vecta 46 group and 17.2 ± 10.1 in the Vecta 46 group. There were no statistically significant differences in patient sex, comorbidities, or baseline presentation between the non-Vecta 46 and Vecta 46 groups. These baseline characteristics and significant values for differences between groups are listed in Table 1.

**Table 1:** Baseline Characteristics.

|  | Non-Vecta 46 n = 142 | Vecta 46 n = 32 | Total n<br>=174 | p-value |
| --- | --- | --- | --- | --- |
| **Age (years)** | 69.4 ± 14.9 | 68.7 ± 17.8 | 69.47 ±<br>16.0 | 0.537 |
| **Patient Sex** |  |  |  | 0.500 |
| Male | 77 (54.2%) | 16 (50.0%) | 93 |  |
| Female | 65 (46.5%) | 16 (50.0%) | 81 |  |
| **Risk Factors** |  |  |  |  |
| Atrial<br>Fibrillation | 37 (26.1%) | 4 (12.5%) | 41 | 0.161 |
| Diabetes<br>Mellitus | 21 (14.7%) | 7 (21.9%) | 28 | 0.472 |
| Hypertension | 94 (66.2%) | 22 (69.0%) | 116 | 0.945 |
| CAD | 39 (27.5%) | 6 (18.8%) | 45 | 0.427 |
| Previous<br>CVA/TIA | 25 (17.6%) | 5 (15.6%) | 30 | 0.993 |
| Smoking | 44 (31.0%) | 6 (18.8%) | 51 | 0.419 |
| **Baseline mRS** | 0.570 ± 1.01 | 0.813 ± 1.62 | 0.615 ±<br>1.14 | 0.804 |
| **NIHSS on**<br>**Admission** | 17.2<br>± 7.66 | 17.2 ± 10.1 | 17.2 ±<br>8.15 | 0.758 |

### 3.2 Procedure Details

The use of aspiration versus SR differed significantly between the non-Vecta 46 and Vecta 46 groups (p = 0.00325). Aspiration alone was used most frequently in both the Vecta 46 and non-Vecta 46 group and aspiration alone was used in 70 (49.3%) non-Vecta 46 MTs and in 18 (56.3%) Vecta 46 MTs. Aspiration failed and was rescued with SRs in 25 (17.6%) non-Vecta 46 MTs and 6 (18.8%) Vecta 46 MTs. SR alone was used in 37 (26.1%) non-Vecta 46 MTs and 4 (12.5%) Vecta 46 MTs. SRs failed and required aspiration rescue in 1 (0.7%) non-Vecta 46 MTs and 4 (12.5%) Vecta 46 MTs. Other techniques were used in 12 (8.2%) non-Vecta 46 MTs and 0 Vecta 46 MTs. There was a statistically significant difference in number of aspiration attempts with 1.12 ± 0.650 attempts in the non-Vecta 46 group and 1.66 ± 0.963 attempts in the Vecta 46 group (p = 0.00135). The number of SR attempts did not differ significantly between the groups, with 1.38 ± 1.23 attempts in the non-Vecta 46 group and 1.79 ± 2.15 attempts in the Vecta 46 group (p = 0.866). These results are listed in Table 2.

**Table 2:**
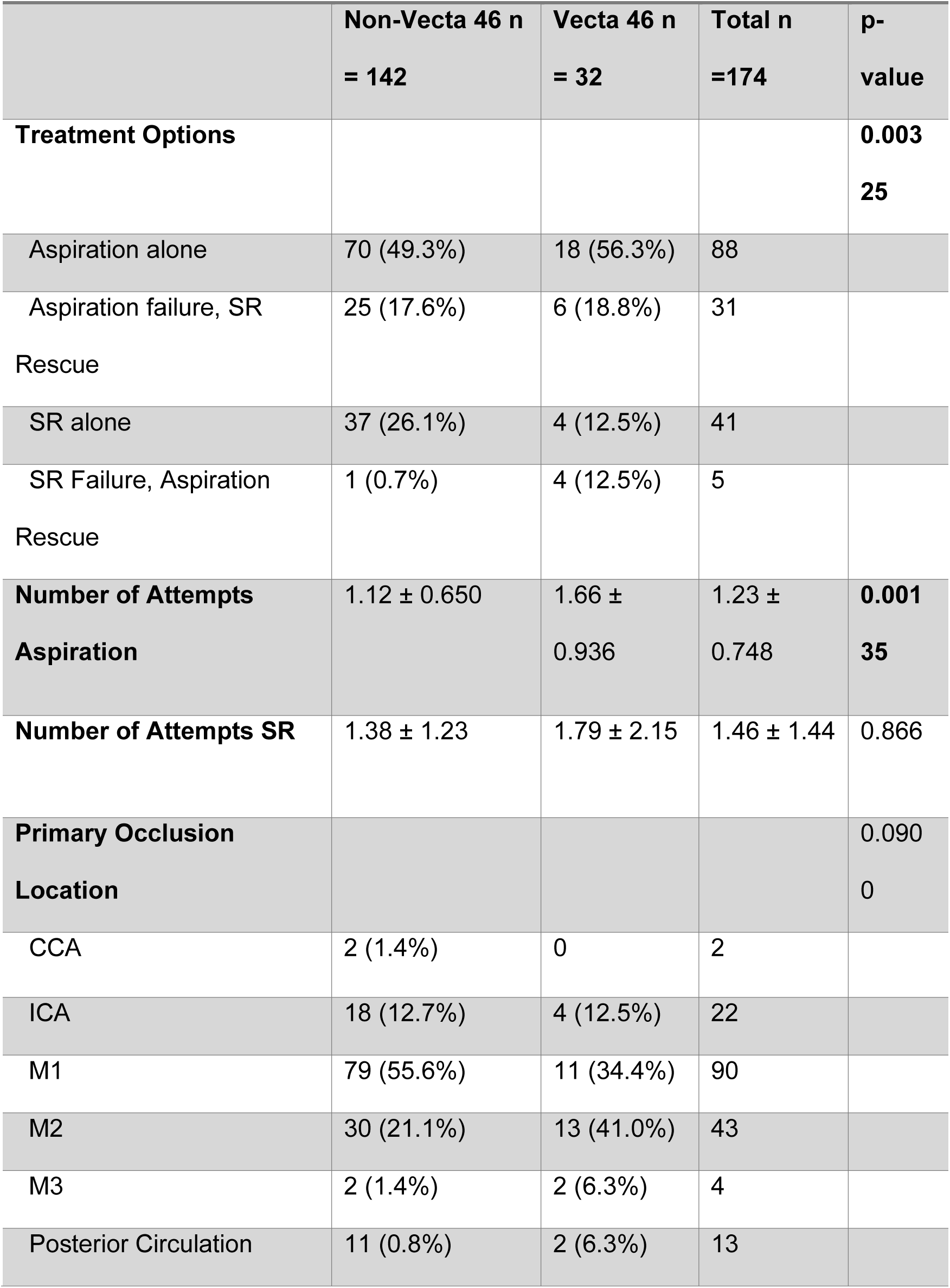
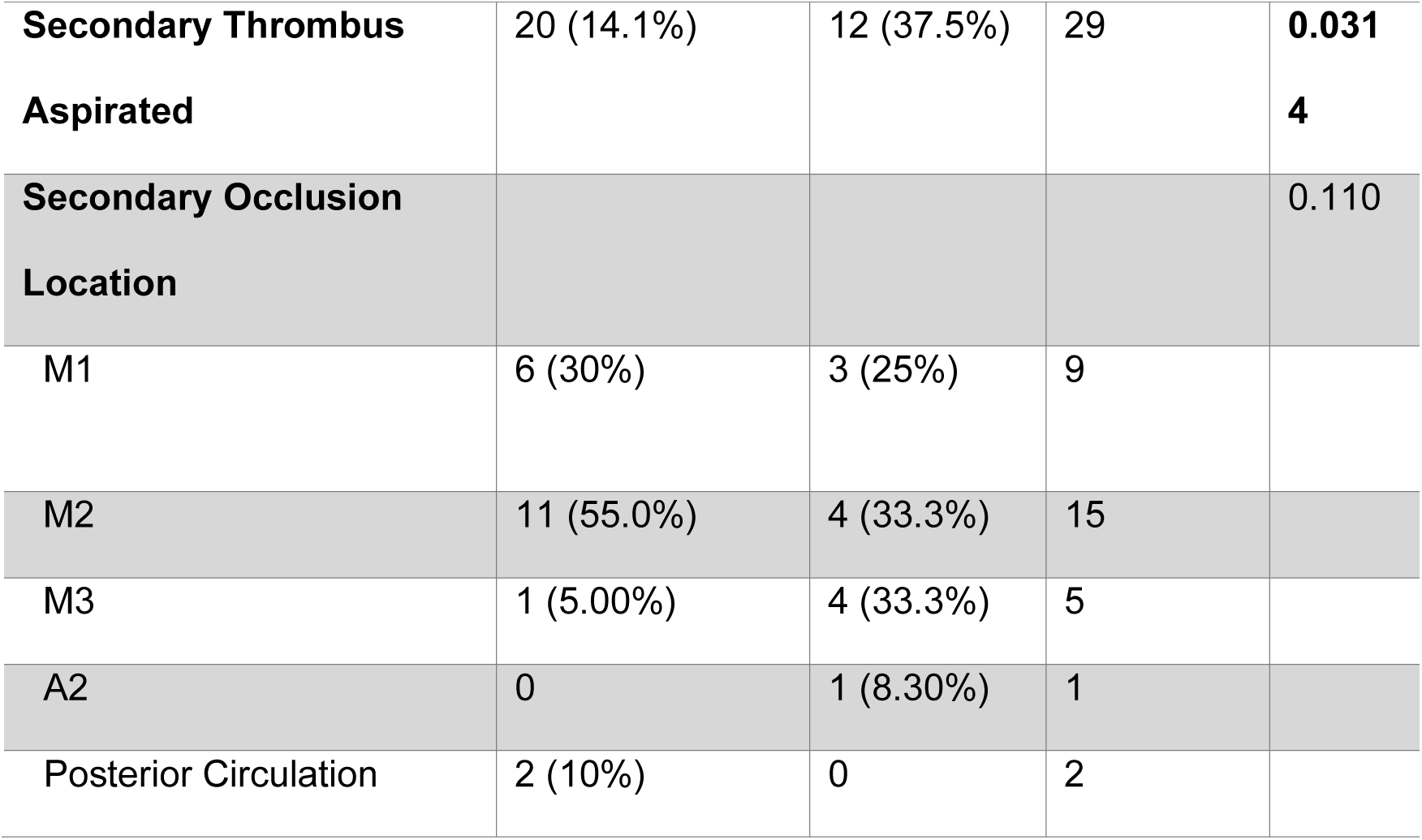
Procedure Details.

The rate of secondary thrombus aspiration differed significantly between the non-Vecta 46 and Vecta 46 groups, with 20 (14.1%) secondary aspirations in the non-Vecta 46 group and 12 (37.5%) secondary aspirations in the Vecta 46 group. There was no statistically significant difference in location of primary (p = 0.09) or secondary (p = 0.11) occlusion aspirated between the non-Vecta 46 and Vecta 46 groups. Figure 1 displays the distribution of primary and secondary occlusion locations in the non-Vecta 46 and Vecta 46 groups. Primary occlusions aspirated in the non-Vecta 46 group were M1 > M2 > ICA > posterior circulation > CCA = M3 and M2 > M1 > ICA > M3 = posterior circulation in the Vecta 46 group (Fig 1A). Secondary occlusions aspirated in the non-Vecta 46 group were M2 > M1 > posterior circulation > M3 and M2 = M3 > M1 > A2 (Fig 1B).

**Figure 1:**
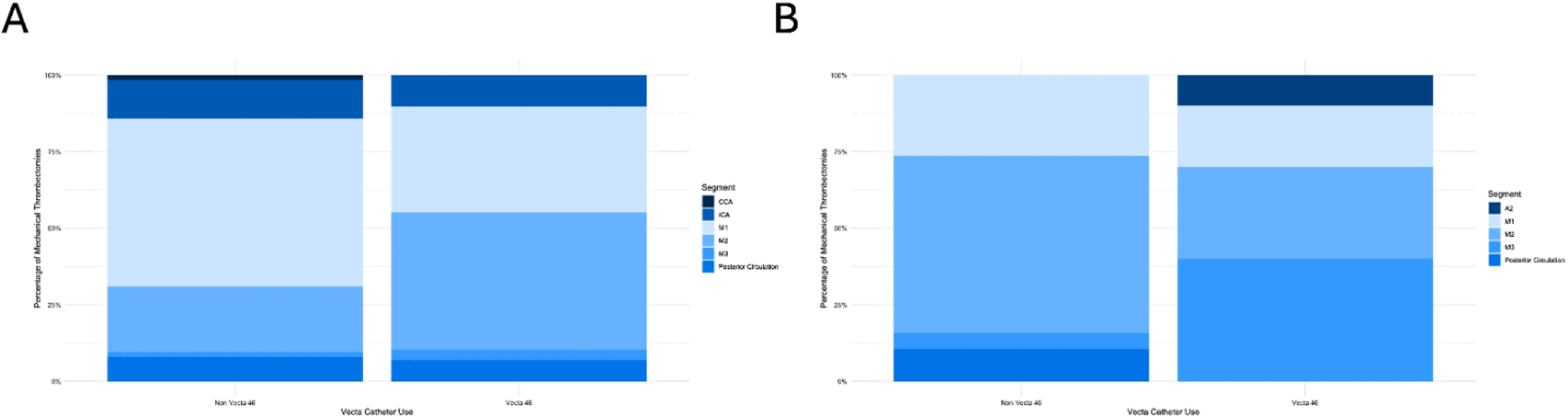
Distribution of (A) primary thrombi and (B) secondary thrombi aspirated in the non-Vecta 46 vs Vecta 46 group.

### 3.3 Outcomes

Table 3 lists procedure outcomes in the Vecta 46 or non-Vecta 46 group. Procedure outcomes did not differ significantly between groups. Successful canalization occurred in 141 (99.3%) patients in the non-Vecta 46 cohort and 31 (96.9%) patients in the Vecta 46 cohort (p = 0.800). Time to recanalization was 23.1 ± 20.8 minutes in the non-Vecta 46 cohort and 22.6 ± 11.1 minutes in the Vecta 46 cohort (p = 0.245). Procedure duration was 57.8 ± 43.92 in the non-Vecta 46 group and 47.6 ± 26.4 in the Vecta 46 group (p = 0.580). Discharge mRS score was 2.82 ± 2.06 in the non-Vecta 46 group and 2.81 ± 2.16 in the Vecta 46 group (p = 0.875).

**Table 3:**
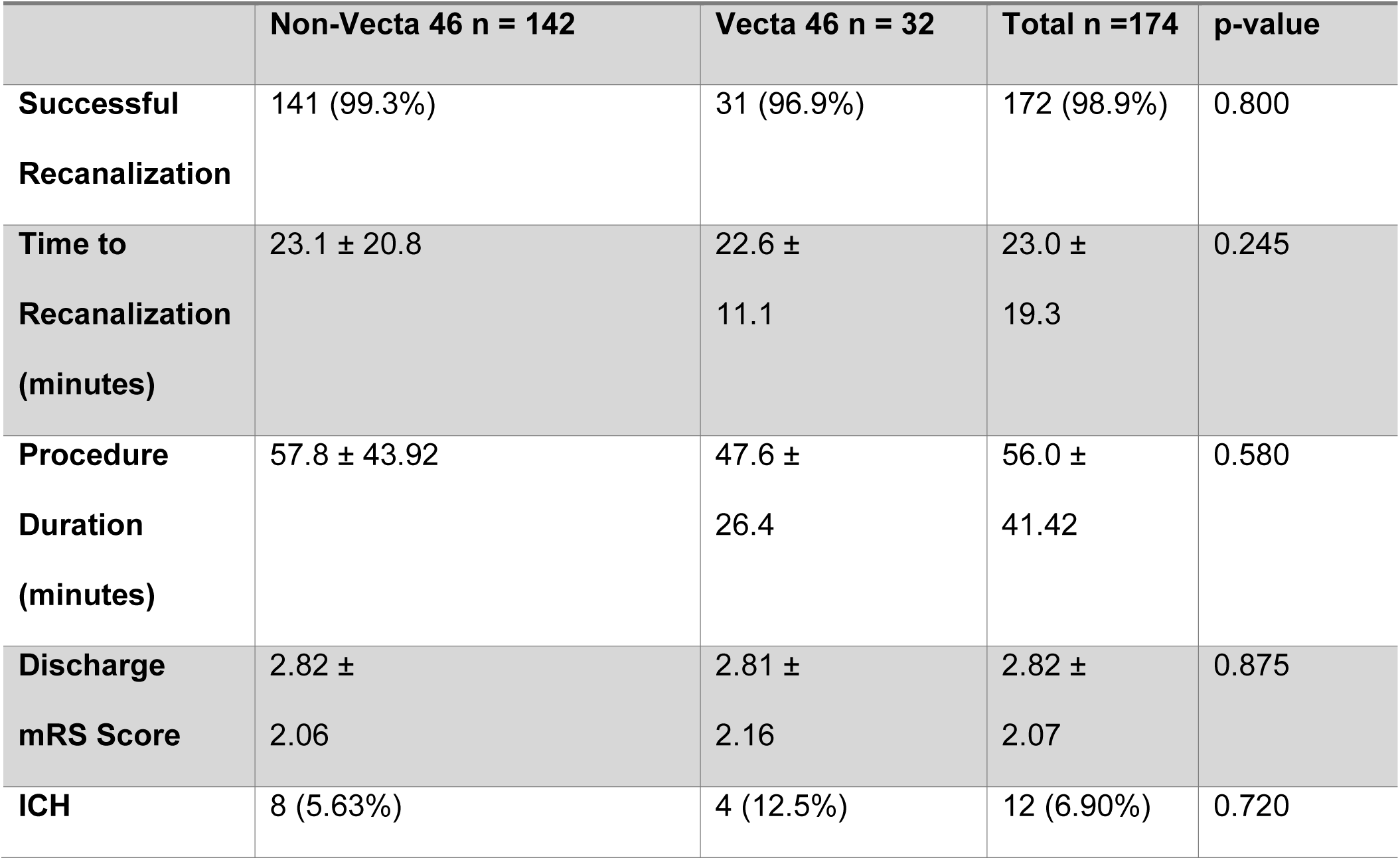
Outcomes.

Hemorrhage occurred in 8 (5.63%) patients in the non-Vecta 46 group and 4 (12.5%) patients in the Vecta 46 group (p = 0.720).

## 6. Discussion

This single-center retrospective investigation compared the safety and efficacy profile of the Vecta 46 intermediate catheter in MT of LVOs and MeVOs to MTs performed with all other catheter configurations. Distal access catheters are growing in popularity for MT as they provide rapid, stable intracranial access.^16^ As new devices, such as the Vecta 46, come to market, it is important to compare their safety and efficacy to established options. There were no statistically significant differences in revascularization time, intracranial hemorrhage, or functional outcome between Vecta 46 and other catheter configurations. These results agree with non-inferiority of distal access catheters to stent retrievers and aspiration catheters reported in the literature.^17^ As well as non-inferiority between the larger Vecta 71 and HiPoint Reperfusion distal catheters.^18^ Given this parity, our results support the Vecta 46 as a viable option to be chosen based on anatomy and operator preference, as supported by the American Heart Association Scientific Statement.^19^

Revascularization time is critical in acute ischemic strokes.^20^ Every 15-minute delay in revascularization time significantly increases the likelihood of symptomatic intracranial hemorrhage, failure to discharge home, and in-hospital mortality.^21^ There were no statistically significant differences in revascularization time between Vecta 46 and other catheter configurations, the primary endpoint of our investigation. Patients treated with the Vecta 46 also had similar functional outcomes to other catheter configurations, a reflection of non-inferior time to revascularization.^22^

Despite no difference in revascularization and functional outcome, Vecta 46 thrombectomies had a unique profile of treatment compared to other options in our cohort, initiating MT with direct aspiration and failing attempts initiating with SRs more often. This likely reflects the intended design of the Vecta 46 for direct aspiration and operator experience with Vecta 46’s significantly higher SR failure rate. This is the first report of the poor efficacy of preliminary SR use with Vecta 46, an important consideration when selecting catheter system if anatomy is unfavorable for aspiration.^23^ Another important consideration in use of the Vecta 46 is a requirement for more attempts at direct aspiration to achieve reperfusion. Compared to thrombectomies with other catheter systems, it was more likely to require 2, 3, or 4 attempts at direct aspiration. A similar trend is seen in the larger sized Vecta 71, indicating possible inferiority in achieving first-pass revascularization throughout the Vecta group of catheters.^18^ This may be a reflection of the Vecta 46’s smaller size, as decreasing catheter size reduces rate of first-pass recanalization, as aspiration force scales with diameter.^24^ Overall, we have elucidated a significant difference in procedure characteristics but also demonstrated non-inferior safety or efficacy outcomes for the Vecta 46, reflecting changing practices in thrombectomies to prioritize faster access while sacrificing procedure brevity.

The strength of this investigation is the breadth of procedure details, occlusion types, and a comparison to thrombectomies completed by the same operators. Current literature on the Vecta 46 is minimal, only including MeVO thrombectomies and lacking a control group, making it poorly generalizable.^15^ Our results provide a broader investigation of the full spectrum of indications for Vecta 46, including direct aspiration and SR MT for LVOs and MeVOs. Although there were no statistically significant differences in primary or secondary occlusion location between Vecta 46 and non-Vecta 46 thrombectomies, Vecta 46 was used to aspirate secondary occlusions more often, likely due to versatility and trackability in navigating distal vasculature while retaining sufficient aspiration force to provide good results for thrombectomy. A direct comparison in another study is needed to determine if passes previously attempted with SR rescues can be replaced with aspiration attempts when using intermediate catheters, given the Vecta 46’s inferiority in SR rescue.

A limitation of this investigation is it is a single-center, retrospective study, which requires careful consideration before integration into wider clinical practice. Although baseline patient characteristics were similar, more secondary occlusions were aspirated with the Vecta 46, meaning the reported outcomes may be skewed by analysis of two different patient populations. Randomized controlled trials and meta-analyses agree with our results,^25,26^ strengthening our conclusions, several randomized controlled trials comparing the efficacy of Vecta 46 to other aspiration and SR options in both LVOs and MeVOs are needed. We emphasize that our analysis is not a clinical trial; rather, an observational comparison intended to demonstrate that the introduction of a novel intermediate catheter has yielded outcomes in line with our historical experience using standard devices, and to further provide context for the expanding role of MT in MeVO stroke. By sharing our single-center experience, we aim to contribute needed clinical data on the feasibility of using an intermediate-caliber aspiration catheter for both LVO and MeVO MT, as such data are crucial for guiding device selection and supporting the safe expansion of MT to MeVOs. This may provide further impetus for evaluating new device innovations in both distal and proximal occlusion MTs.

Overall, the Vecta 46 intermediate catheter is a versatile option for endovascular treatment of ischemic stroke. At our institution, it is employed for LVOs and MeVOs with similar safety and efficacy to other endovascular treatment options. Compared to other catheters, direct aspiration is more likely to be attempted with the Vecta 46 than SR and more passes are required to achieve revascularization. Despite these procedural differences, there are no differences in time to rate of intracranial hemorrhage, or discharge mRS. Altogether, the Vecta 46 has similar safety and outcomes compared to other endovascular treatment options despite procedural differences.

## Data Availability

Deidentified data available on request from the authors

## 7. Acknowledgments

The preliminary results of this investigation were presented at the Society for Neurointerventional Surgery 2025 annual conference.

## 8. Sources of Funding

None

## 9. Discosures

None.

## Notes

### Competing Interest Statement

The authors have declared no competing interest.

### Clinical Trial

This research received no specific grant from any funding agency in the public, commercial, or not-for-profit sectors.

### Funding Statement

The authors report no funding source.

### Author Declarations

Investigation approved by the University of Florida institutional review board (IRB202400732).

